# Crystal Clear: Purity of consumer-level methamphetamine samples and methamphetamine-adulteration of other drugs in Los Angeles, 2023-2025

**DOI:** 10.1101/2025.03.18.25323868

**Authors:** Adam J. Koncsol, Caitlin A. Molina, Ruby Romero, Megan Grabill, Morgan E. Godvin, Jesse Carrizal, Manuel Contreras, Oscar Arellano, Naomi Villanueva, Elham Jalayer, spider davila, Soma Snakeoil, Joshua Smith, Charles J. Santamaria, Leslie Nuñez, Ezinne Okonkwo, Siddarth Puri, Joseph R. Friedman, Chelsea L. Shover

## Abstract

**Introduction:** The United States (US) has seen a rise in methamphetamine use disorders and methamphetamine-involved deaths. The US Department of Justice and Drug Enforcement Agency reports that seized wholesale methamphetamine is almost uniformly high purity. However, there is limited information on the concentration of methamphetamine at the consumer level. We leverage data from a community-based drug checking program in Los Angeles to investigate (1) the makeup of the retail illicit methamphetamine market and (2) the extent to which methamphetamine plays a role as an adulterant in other drugs.

**Methods:** Anonymous participants accessing a community-based drug checking program voluntarily provided samples of illicit drug products and completed brief interviews at four sites in Los Angeles County, California, from February 2023 to January 2026. Samples underwent laboratory-based qualitative testing via direct analysis in real-time mass spectrometry (DART-MS) and quantitative liquid-chromatography mass spectrometry (LC/MS). We quantitated the presence and concentration of methamphetamine as well as all other identified substances among 1) samples expected only to be methamphetamine and 2) samples expected to be another substance (e.g., fentanyl, heroin) where methamphetamine was found as an adulterant/contaminant.

**Results:** Of N=2193 total samples, n=652 (29.7%) were methamphetamine-positive on DART-MS. Among methamphetamine-positive samples, the drug was an expected component in two thirds of samples and unexpected in one third. In samples expected to contain methamphetamine, the mean purity was 78.2% (SD 24.0%; range <1% to ≈100%). As an unexpected component, the mean methamphetamine purity was 17.8% (SD 32.3%), and was mostly commonly found in samples expected to be fentanyl, heroin, MDMA, other amphetamines (e.g., counterfeit Adderall), and 2C-B.

**Conclusions:** Consumers in Los Angeles seeking methamphetamine appear to frequently obtain high-purity samples that are rarely adulterated with other substances, although we observed a wide variation in concentration between samples. In contrast, drugs purchased without the expectation of containing methamphetamine had substantively high prevalence of methamphetamine adulteration/contamination. Both intentional and unwitting consumers may be vulnerable to the health risks of potent and variable methamphetamine, including psychiatric, cardiovascular, and other illnesses. Future research should expand methamphetamine surveillance techniques and investigate how variability in illicit markets may impact health outcomes.

**Highlights:**

- Samples expected to contain methamphetamine had a mean purity of 72% (SD 24%).
- For 90% of samples expected to be meth, no other active compounds were detected.
- Meth was a substantial adulterant in other drugs, e.g. opioids (mean purity 17%).

## Introduction

Methamphetamine is a central nervous system stimulant that carries potential health risks to multiple organ systems and is implicated in a rising number of drug-related deaths ^1–5^. Methamphetamine use appears to be increasing around the world, particularly in the Americas, Asia, and Australia ^6–9^. Health consequences of methamphetamine use have also increased, with the U.S. seeing a rise in methamphetamine-related deaths from 4,500 in 2015 to approximately 30,000 in 2024 (National Center for Health Statistics, 2025). Chronic methamphetamine use increases the risk of cardiovascular harms such as myocardial toxicity, coronary artery disease, hypertension, stroke, sudden cardiac arrest, and other causes of morbidity and mortality ^1–5^.

As a fully synthetic drug, methamphetamine is considerably cheaper and easier to produce than agriculturally-derived stimulants like cocaine ^10–12^. Methamphetamine seized by law enforcement tends to have exceptionally high purity, with most recently available U.S. national averages exceeding 90% ^13–15^. However, drugs seized in criminal investigations are potentially interdicted higher in the supply chain prior to adulteration by lower-level sellers and may reflect purer product than what end-consumers receive. Drug checking—chemically testing personal samples of drugs for harm reduction and supply monitoring—can characterize methamphetamine purity and adulteration at the consumer level.

Los Angeles County, California has long led the United States in both number of methamphetamine-related deaths (reflecting the large population) and proportion of all drug-related deaths attributable to methamphetamine (reflecting a persistent market) ^16^. In recent years, an increasing number of Los Angeles County’s deaths attributed to methamphetamine were also attributed to fentanyl ^16^. This pattern is mirrored in national data, where the fourth wave of the U.S. overdose crisis is characterized by deaths involving polysubstance combinations of fentanyl and other synthetic opioids and stimulants like cocaine ^17,18^. While public health campaigns have often focused on the threat of fentanyl adulteration of other drugs, data from testing actual drug products has been limited. Leveraging data from a community-based drug checking program, we sought to empirically investigate the methamphetamine supply in Los Angeles. Specifically, we sought to characterize the presence and purity/concentration of methamphetamine and co-detected substances among 1) methamphetamine-positive samples expected only to be methamphetamine and 2) methamphetamine-positive samples expected to be another substance (e.g., purchased as fentanyl or heroin) where methamphetamine is functioning as an adulterant. “Purity” is used to refer specifically to the percentage by mass of methamphetamine in samples expected to be methamphetamine, whereas “concentration” is used to refer to the percentage by mass of any compound present in any drug (e.g., methamphetamine in a sample expected to be fentanyl).

## Methods

Anonymous participants utilizing a community-based drug checking program, *Drug Checking Los Angeles*, voluntarily provided a small sample (approximately 1-5 mg) of drug products (e.g., powders, crystals, pills, liquids, tar, etc.) for testing at four different sites in Los Angeles County, California: (1) East Los Angeles, (2) Downtown Los Angeles / Skid Row, (3) Hollywood, and (4) South Los Angeles. This study uses data collected from February 2023 to December 2025. An optional survey was administered by trained staff to assess what the drug was purported to be at the time of sale and what a participant expected the sample to contain. Instances where a respondent did not know what the sample contained or declined to answer are presented as “Unknown / Declined.”

Initial-point-of-service drug checking used Fourier-Transform Infrared (FTIR) spectroscopy and various BTNX-branded and lab-validated immunoassay test strips, such as fentanyl and methamphetamine test strips ^19^. Samples received through Drug Checking Los Angeles were tested in the field to provide real-time results to participants and also sent by mail to the National Institute of Standards and Technology Rapid Drug Analysis and Research (NIST RaDAR) program for laboratory-based analysis ^20^. NIST RaDAR evaluated all samples included in the study with direct analysis in real time mass spectrometry (DART-MS) and a subset of samples with liquid-chromatography mass spectrometry (LC/MS).

DART-MS methods assess samples against a library of nearly 1,500 substances, including most pharmaceutical and illicitly manufactured drugs, adulterants, cutting and bulking agents, precursor chemicals, and other relevant substances (adhesives, industrial ingredients, etc.). DART-MS methods are only able to provide information on the binary presence or absence of compounds and are unable to provide information on concentration; however, they only require residual or trace amounts of a sample to be analyzed. A subset of samples analyzed with DART-MS were also analyzed with LC/MS methods to quantitate the percent mass of pre-specified compounds present in the sample. See **Supplemental Table A** for more information about the specific analytes used on the LC/MS quantitation panel. More detailed descriptions of the confirmatory DART-MS and LC/MS methodologies employed in this study have been previously published ^21^. An important limitation to all reported results is that laboratory methods may be unequipped to detect byproduct or precursor compounds related to the synthesis products for which laboratory standards do not exist.

Methamphetamine concentration values reported by NIST RaDAR were expressed as methamphetamine freebase. To account for the molecular weight of the hydrochloride salt when calculating concentration estimates, methamphetamine freebase was converted to methamphetamine HCl. Dividing the molecular weight of methamphetamine HCl (185 Da) by the molecular weight of methamphetamine freebase (149 Da) and multiplying that conversion factor (approximately 1.24) by each methamphetamine freebase value can provide a more accurate estimation of methamphetamine concentration in our sample—especially because all samples of methamphetamine tested were sold as “crystal methamphetamine” or “ice”. As a result of the freebase-to-salt concentration conversion, any samples ≥85% methamphetamine freebase were treated as pure and were imputed to be 100% for this analysis. Similarly, samples that were considered below the limit of quantitation (<LOQ) were imputed to be 0.1% to calculate descriptive statistics.

The count and prevalence (%) of any co-detected compounds were calculated across methamphetamine-expected and methamphetamine-unexpected samples confirmed with DART-MS to contain methamphetamine. This highlights (1) compounds that may be present in samples expected to be methamphetamine and (2) how often methamphetamine was detected in samples expected to be other drugs, like heroin or fentanyl. For the subset of samples that underwent quantitative LC/MS analysis, the concentration was assessed in terms of average, standard deviation, and range of methamphetamine, presented across participant drug expectation categories. Study protocols were approved by the UCLA IRB (IRB-22-0760 and IRB-22-1273). FTIR analyses were completed using OPUS, version 8.7.31 (Bruker, Billerica, MA, USA). Statistical analyses were carried out using Stata, version 19.5 (StataCorp, College Station, TX, USA).

## Results

We tested N = 2,193 samples of drug product with DART-MS from February 2023 to December 2025. Among n = 652 samples found to contain methamphetamine, 67% were expected to contain methamphetamine (n = 432 methamphetamine-only, n = 2 methamphetamine + heroin mixture, n = 1 methamphetamine + fentanyl mixture, and n = 1 methamphetamine + ketamine mixture). The remaining 33% (n=220) were not expected to contain methamphetamine.

**Table 1**. shows co-detected substances among n=432 of methamphetamine-positive samples expected to only contain methamphetamine. The most commonly co-prevalent compounds were cocaine (n=9, 2.1%), ephedrine (n=8, 1.9%), lidocaine (n=8, 1.9%), and fentanyl (n=7, 1.6%). Most samples (n=387, 89.6%) expected to be methamphetamine contained only methamphetamine, with no other co-prevalent compounds detected on DART-MS.

Of the n=652 samples confirmed to contain methamphetamine, a subset (n=208) underwent LC/MS analysis. For quantitated samples expected to be methamphetamine (n=172), the average percent by mass of methamphetamine hydrochloride was 78.2% (SD=24.0%) and ranged from below the limit of quantitation (≈ 0.1%) to virtually pure (≈100%). **Figure 1** highlights the distribution of methamphetamine concentration among samples expected to be methamphetamine vs where methamphetamine was not expected, excluding n=11 samples where the expected drug could not be determined. For samples which were not pure methamphetamine but did not contain any additional compounds on DART-MS, the remaining percentage mass of the sample was likely to be inert precursors and byproducts related to inefficient methamphetamine synthesis.

**Table 1.**
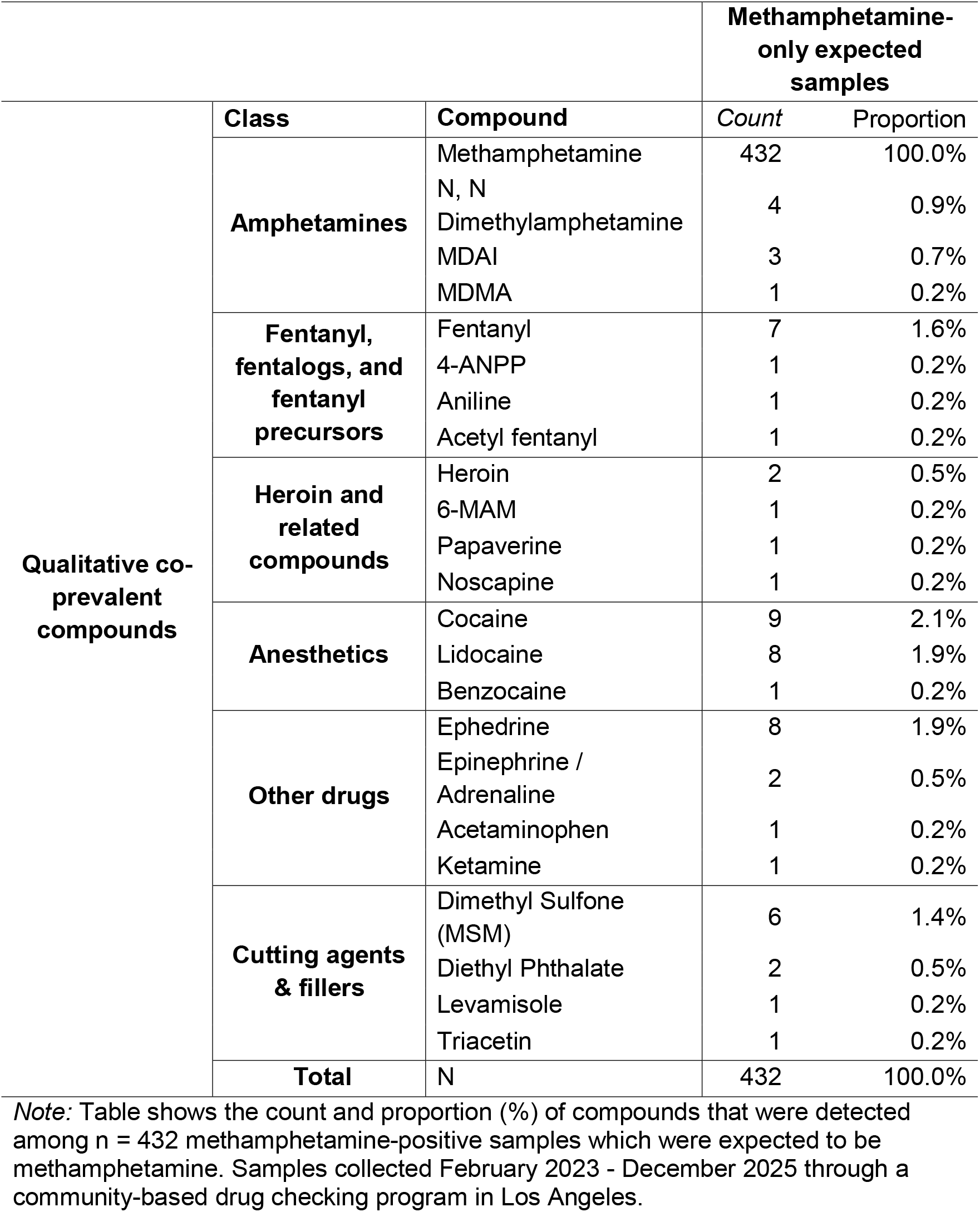
The count and proportion of prevalent compounds among only-methamphetamine expected and positive samples.

**Fig. 1.**
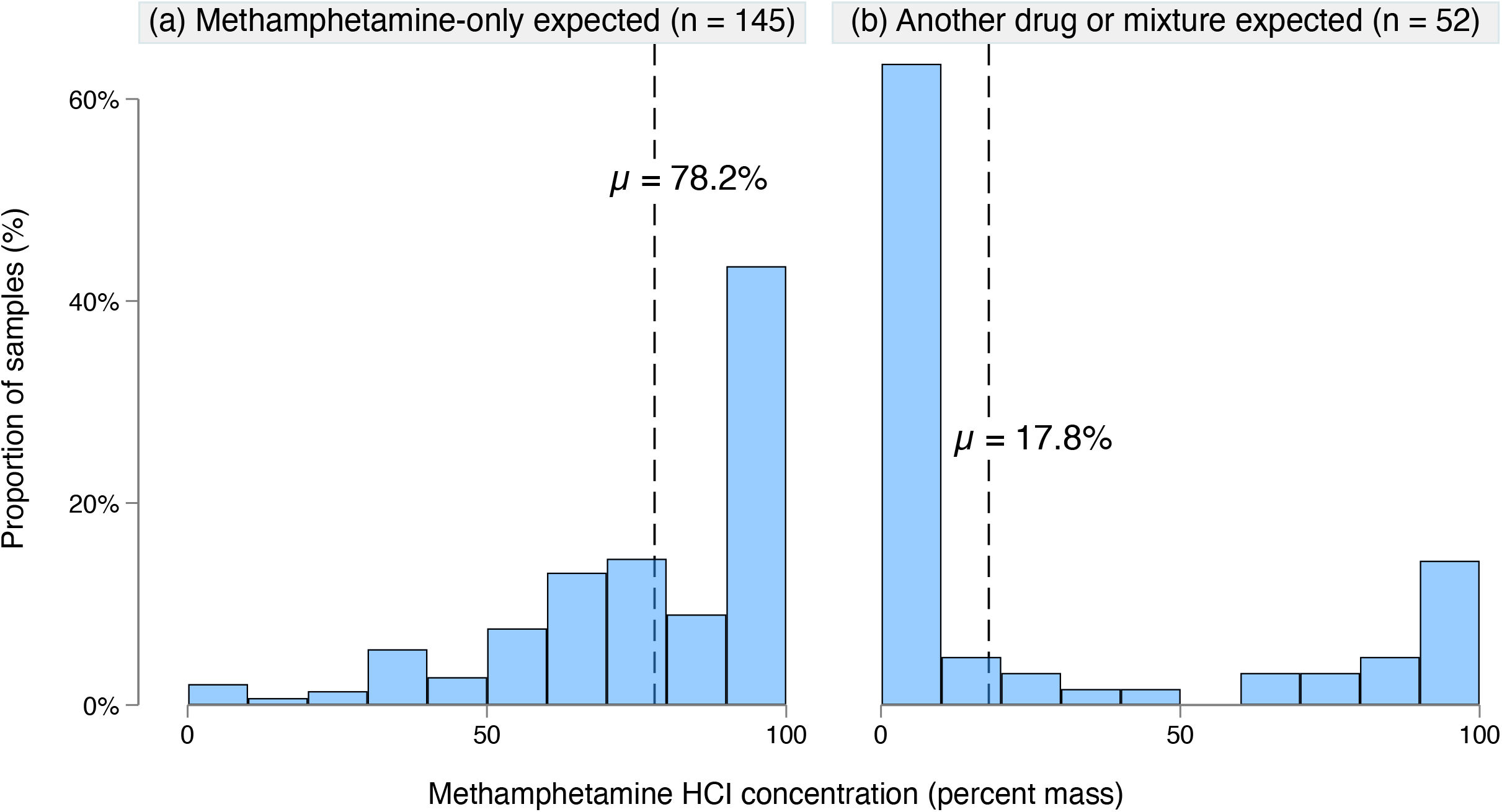
Methamphetamine concentration among samples (a) expected to methamphetamine or (b) another expected drug. Note: Graph shows distribution of methamphetamine concentration among samples expected to be methamphetamine and samples expected to be another drug or mixture (unknown expectation excluded). N= 197 methamphetamine-positive LCMS-quantitated samples collected Feb. 2023 - Dec. 2025.

In approximately 33% (n=220) of samples in which methamphetamine was detected, its presence was not expected by the participant. **Table 2 and Figure 2** highlight how frequently methamphetamine was detected as an adulterant in drug products expected to be other substances. Notably, methamphetamine was detected in 8.5% of expected fentanyl samples (n=62), 16.4% of expected heroin samples (n = 34), 14.0% of expected MDMA / MDA samples (n =19), and 70.4% of expected amphetamine samples (n =19). It was also detected among samples expected to be 2C-B (41.2%, n=7), benzodiazepines (11.5%, n = 6), and GHB / BDO (33.3%, n=2). In our sample, methamphetamine was occasionally detected in samples expected to be oxycodone (4.0%, n =3), powder and crack cocaine (3.8%, n=6), and was not detected in the few samples expected to be designer stimulants, cathinones, or research chemicals others brought for testing. Although we are cautious to extrapolate due to small sample sizes, these emerging compounds may be ripe for methamphetamine adulteration given their entactogenic and stimulating effects as well as relatively significantly higher market prices.

**Table 2.** Methamphetamine positivity by drug expectation.

| **Expected drug** | **Methamphetamine negative** |  | **Methamphetamine positive** |  | **Total samples** |  |
| --- | --- | --- | --- | --- | --- | --- |
|  | *Count* | *Proportion* | *Count* | *Proportion* | *Count* | *Proportion* |
| Methamphetamine | 9 | 2.0% | 432 | 98.0% | 441 | 20.1% |
| Fentanyl | 671 | 91.5% | 62 | 8.5% | 733 | 33.4% |
| Heroin | 173 | 83.6% | 34 | 16.4% | 207 | 9.4% |
| Amphetamine | 8 | 29.6% | 19 | 70.4% | 27 | 1.2% |
| MDMA / XTC | 117 | 86.0% | 19 | 14.0% | 136 | 6.2% |
| 2C-B | 10 | 58.8% | 7 | 41.2% | 17 | 0.8% |
| Benzodiazepines | 46 | 88.5% | 6 | 11.5% | 52 | 2.4% |
| Cocaine or Crack | 152 | 96.2% | 6 | 3.8% | 158 | 7.2% |
| "Psilocybin" products | 5 | 62.5% | 3 | 37.5% | 8 | 0.4% |
| Oxycodone | 72 | 96.0% | 3 | 4.0% | 75 | 3.4% |
| Heroin and Methamphetamine mixture | 0 | 0.0% | 2 | 100.0% | 2 | 0.1% |
| GHB or BDO | 4 | 66.7% | 2 | 33.3% | 6 | 0.3% |
| Amoxicillin | 0 | 0.0% | 1 | 100.0% | 1 | 0.0% |
| Cannabis | 1 | 50.0% | 1 | 50.0% | 2 | 0.1% |
| Dexmethylphenidate | 0 | 0.0% | 1 | 100.0% | 1 | 0.0% |
| Methamphetamine and Fentanyl mixture | 1 | 50.0% | 1 | 50.0% | 2 | 0.1% |
| Methamphetamine and Ketamine mixture | 0 | 0.0% | 1 | 100.0% | 1 | 0.0% |
| Fentanyl and Heroin mixture | 4 | 80.0% | 1 | 20.0% | 5 | 0.2% |
| PCP | 6 | 85.7% | 1 | 14.3% | 7 | 0.3% |
| Ketamine | 78 | 98.7% | 1 | 1.3% | 79 | 3.6% |
| Unknown/declined | 122 | 71.3% | 49 | 28.7% | 171 | 7.8% |
| Total | 1541 | 70.3% | 652 | 29.7% | 2193 | 100.0% |

Note: There were n = 652 samples of drug product that were confirmed positive for methamphetamine and are a subset of N = 2,193 total samples that were brought for testing at a community-based drug checking site in Los Angeles. Samples collected February 2023 - December 2025. DART-MS methods were used to qualitatively assess the presence of methamphetamine. Semi-structured ethnographic interviewing methods were used to capture what participants expected their drugs to be at the time of testing, either through what the substance was bought as or what the participant otherwise expected the sample to contain and may be subject to bias.

**Fig 2.**
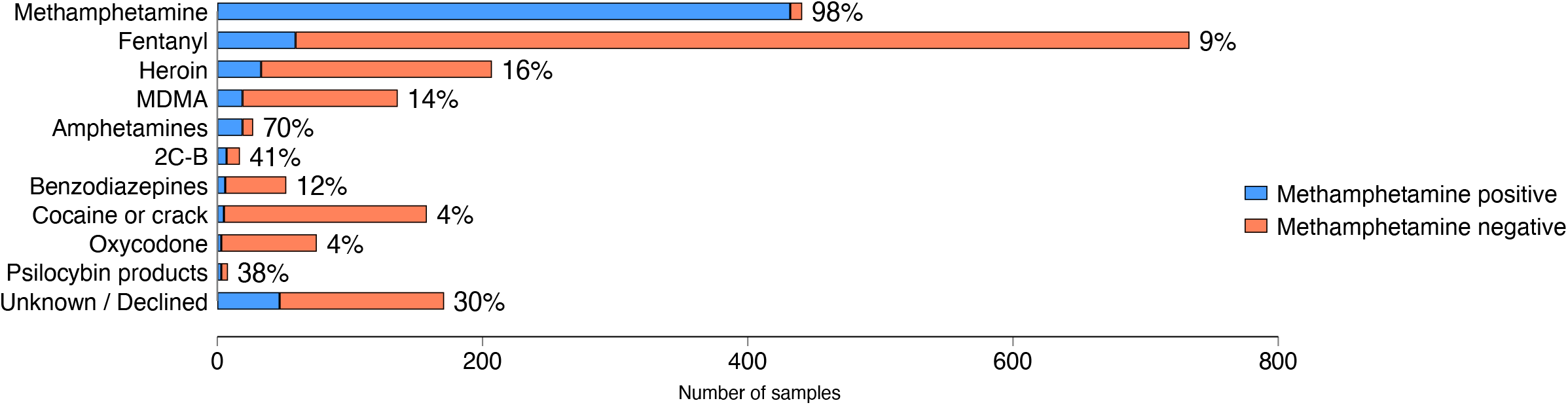
Prevalence of Methamphetamine Detection Across Expected Drug Categories. Note: Graph shows methamphetamine positivity among different expected drugs. Bar percentages represent the proportion of samples positive for methamphetamine relative to all samples, per category. N = 2,193 total samples of drug product analyzed, with methamphetamine detected in n = 652. Data collected February 2023 - December 2025 in Los Angeles.

**Table 3** displays the concentration of methamphetamine in samples expected to be methamphetamine, as well as its concentration in other drugs (when it is present as an adulterant). When present as an adulterant in other drugs, methamphetamine was present at levels that are likely clinically relevant. For example, among 30 samples expected to only contain fentanyl, the mean methamphetamine concentration was 17.5% by mass. Similarly, in 9 samples expected to only contain heroin, the average methamphetamine concentration was 15.2% by mass. Notably, however, was how heterogeneous methamphetamine adulteration was across samples of heroin and fentanyl, with standard deviations of methamphetamine concentration being 33.0% and 27.0%, respectively.

**Table 3.** Methamphetamine appears in other illicit drugs, sometimes in significant quantities.

|  |  | Average<br>methamphetamine concentration<br>(percent by mass) | Standard<br>Deviation | Range | N |
| --- | --- | --- | --- | --- | --- |
| **Expected**<br>**drug or**<br>**substance** | Methamphetamine | 78.2 | 24.0 | [<LOQ -<br>≈100%] | 145 |
|  | Fentanyl | 17.5 | 33.0 | [<LOQ -<br>≈100%] | 30 |
|  | Amphetamines | 19.5 | 32.6 | [<LOQ -<br>90.0%] | 8 |
|  | Heroin | 15.2 | 27.0 | [<LOQ -<br>81.3%] | 9 |
|  | 2C-B | 4.9 | . | 4.9% | 1 |
|  | Cocaine / crack | 49.3 | 69.6 | [<LOQ -<br>98.6%] | 2 |
|  | GHB | <LOQ | . | <LOQ | 1 |
|  | Oxycodone | 3.8 | . | 3.8% | 1 |
|  | Unknown /<br>Declined | 65.9 | 38.3 | [24.8% -<br>≈100%] | 11 |
|  | All<br>methamphetamine-positive samples | 62.4 | 37.5 | [<LOQ -<br>≈100.0%] | 208 |

Note: Table shows the methamphetamine concentration (percent by mass), standard deviation, range, count of samples and participant's expected drug or substance among n = 208 methamphetamine-positive, LC-MS quantitated samples. Data collected February 2023 - December 2025 in Los Angeles. Semi-structured ethnographic interviewing methods were used to capture what participants expected their sample to be at the time of testing, either through what the drug or substance was bought as or what the participant otherwise expected the sample to contain. As with other forms of self-reported data, these data may be subject to bias if a
participant mixed up samples or otherwise made an error in recalling what they bought a sample as.

## Discussion

In this study of consumer-level methamphetamine-involved samples, we found that drugs expected to be methamphetamine overwhelmingly—90% of the time— contained only methamphetamine. In contrast, samples expected to only contain other drugs (including fentanyl, heroin, MDMA, 2C-B, and others) often unexpectedly contained methamphetamine. Overall, this reflects the increasingly ubiquitous role of inexpensive and highly pure methamphetamine in various aspects of the illicit drug market, as well as the volatility of the drug supply in the age of synthetic drugs more generally. In our sample, the percentage mass of methamphetamine in a sample expected to be methamphetamine ranged from well below 1% to nearly 100%, with an average of 78%. These findings illustrate that the purity of methamphetamine at the consumer-level in Los Angeles is not as uniform or high as law enforcement seizure data suggest (95%-100%).

Variation in methamphetamine purity may have implications for the health risks of individuals exposed to the drug. Individuals expecting or accustomed to lower purity samples may be inadvertently and unwittingly exposed to a higher dose. This likely has implications for methamphetamine toxicity, including risk of psychosis, mood dysregulation, cardiovascular and pulmonary harms. Similarly, understanding variation in potency may have implications for treatment initiation and retention with medications for methamphetamine use disorder, as well as psychosocial and behavioral interventions such as contingency management, cognitive behavioral therapy, motivational interviewing, and community reinforcement approaches ^22–30^. Understanding methamphetamine purity at the consumer level can help more fully contextualize health-related harms associated with methamphetamine, along with other factors relevant to clinical care, like the dosage, frequency of use, route of administration. Additional public health interventions that include harm reduction programming for people who use methamphetamine may be able to consider that methamphetamine purity varies significantly between batches when suggesting methods to avoid ‘overamping’ and other negative effects of methamphetamine use.

Our findings do not support the notion that fentanyl has been regularly added to drug products sold as methamphetamine in Los Angeles. Fentanyl was detected in 1.6% of samples expected to be methamphetamine, and in the few cases where quantitative data were available, the concentrations were very low. It is unknown how many of these samples were purchased already contaminated versus being later contaminated by co-use through shared paraphernalia or storage, given that polysubstance use was commonly reported by drug checking participants. These findings may not generalize more broadly as drug markets are often highly localized and may change rapidly.

The balance between high-risk yet low probability of fentanyl adulteration in methamphetamine can inform measured public health communications. Any fentanyl cross-contamination, even trace amounts, presents a substantial risk of fatal opioid overdose to people without opioid tolerance; thus, recommendation to check stimulants for fentanyl before use remains an important harm reduction strategy. However, messaging should take care not to imply or assert that methamphetamine adulteration with fentanyl is widespread. Evidence-based messaging can help promote safer practices while also maintaining credibility with people who use drugs and their communities.

Nevertheless, we did find that methamphetamine was regularly detected in samples expected to be other drugs, mostly fentanyl, heroin, and MDMA, sometimes at substantial and potentially clinically relevant amounts. For example, one sample of powder heroin contained 10 times more methamphetamine than heroin by weight: 81% methamphetamine to only 7% of heroin. **These findings help provide context to the role that methamphetamine often can serve in Los Angeles markets: an adulterant to other substances rather than the target of adulteration itself**. Importantly, people who use drugs such as fentanyl, heroin, MDMA, and other amphetamines may unknowingly be exposed to methamphetamine, which could have harmful or unpleasant consequences. When methamphetamine was detected in other drugs, it was typically detected at levels which may be pharmacologically significant. Unintentional methamphetamine exposure may contribute to physical health and psychiatric signs and symptoms inconsistent with the clinical presentation typical of using opioids or shorter-acting stimulants (such as cocaine or MDMA) alone. Harm reduction interventions could underscore the risk of methamphetamine adulteration in other drugs and the potential harms of unintentional exposure. Test strips are one example of a validated, low-cost, and low-barrier technology that could be leveraged to help people avoid unwanted exposure to methamphetamine ^19^.

These findings also help to highlight the potential value of data from community-based drug checking programs. Given that illicit markets are highly volatile and constantly shifting, community-based drug checking programs empower people who use drugs to make decisions in real time while also providing aggregated epidemiological surveillance of multiple, and sometimes overlapping, drug supplies.

### Limitations

While methamphetamine samples tested through a community-based drug checking program can provide unique insights into consumer-level drug markets, there are several limitations to consider. Given the sensitivity of laboratory instruments, detection of adulterants at low levels could occur due to brief, accidental cross-contamination—especially in instances where a person reports that they use both substances. Cross-contamination could also occur through storing multiple drug products in the same bag, or even by touching one substance and then touching another. Although polysubstance use and co-storage information was ascertained by drug checking staff if a participant opted to share that information, it was not possible to completely distinguish between intentional adulteration and accidental cross-contamination. The use of quantitative testing, however, helps to contextualize instances wherein methamphetamine adulteration represents a large portion of the sample tested—far above levels that would be expected from accidental cross-contamination alone.

The fallibility of a participant’s memory to accurately recall drug expectation may have also biased our results. Given that drug expectation was ascertained through participant self-report, a participant may have misremembered or mixed up bags. Despite our best efforts to sample across four geographically and culturally distinct areas of Los Angeles, this is ultimately a convenience sample that may or may not be generalizable to drugs not submitted for checking. For example, a participant may be more likely to bring a sample to check after an adverse or unusual experience or because they were already utilizing SSP services. Laboratory methods are subject to various errors that may affect the precision of our estimates, including the margin of error in quantification and limitations of specific analytical methods. For example, DART-MS techniques are not generally equipped to detect some molecules, like lactose or starches, so the prevalence of some cutting agents, such as lactose, are not reflected in these results. DART-MS methods may have also missed byproduct or precursor compounds related to the methamphetamine synthesis process for which laboratory standards may not yet exist—helping to explain the gap between samples with only methamphetamine detected yet were below 100% pure methamphetamine concentration. The LC/MS quantitation panel only included a subset of all potential compounds, and substances not on this panel were unable to be quantitated. We aimed to mitigate some of these limitations via the use of multiple technologies, yet results should be interpreted in light of these potential sources of error and bias.

## Conclusion

Methamphetamine purity in Los Angeles is high on average, however, lower than the >95% values seen in wholesale seizure data. Few contaminants were detected among samples expected to be methamphetamine; however, the purity of methamphetamine in the community setting is highly heterogeneous between samples. Methamphetamine is frequently used as an adulterant in other drugs—often at pharmacologically-relevant concentrations (i.e., representing a large fraction of the sample). Understanding methamphetamine concentration among drug samples in the community has a number of potential clinical and public health implications. Public health surveillance of consumer-level drug markets may facilitate interventions to address population-wide increases in methamphetamine use, shifting purity, and new adulterants. Future research is needed to evaluate how these findings compare to drug markets beyond Los Angeles. Further research is warranted to understand methamphetamine as an adulterant and the health effects of intentional and unintentional polysubstance use.

**Supplemental table A.** LC/MS Quantitation Panel.

| **Class** | **Quantitative Analyte** |
| --- | --- |
| **Amphetamines** | Methamphetamine |
| **Fentanyl, fentalogs, and fentanyl precursors** | Fentanyl<br>Carfentanil†<br>Fluorofentanyl<br>4-ANPP<br>Ethyl 4-ANPP<br>Phenethyl 4-ANPP |
| **$\alpha_2$- agonists** | Xylazine<br>Medetomidine |
| **Heroin and related compounds** | Heroin<br>6-MAM†<br>Noscapine†<br>Papaverine† |
| **Anesthetics** | Cocaine<br>Lidocaine*<br>Procaine†<br>Tetracaine |
| **Other drugs** | Acetaminophen<br>Caffeine†<br>Ketamine<br>Levamisole†<br>Quetiapine<br>(Seroquel) |
| **Cutting agents & fillers** | BTMPS* |

\*Added to the quantitation panel in July 2024
†Added to the quantitation panel in January 2025

## Data Availability

All data in the present study are not available.

## Funding Acknowledgements

This work was supported by the Centers for Disease Control and Prevention as part of Overdose Data to Action: LOCAL (CDC-RFA-CE-23-0003), National Institutes of Health (R01 DA057630), and an equipment grant from the James B. Pendleton Charitable Trust to the UCLA AIDS Institute and UCLA Center for AIDS Research. AJK was supported by the NIH/National Center for Advancing Translational Science (NCATS) UCLA CTSI (grant TL1TR001883). CLS was supported by the U.S. National Institute on Drug Abuse (grant K01DA050771). The funders had no role in designing the study, analyzing the data, or the decision to submit for publication.

## Additional Acknowledgments

We would also like to acknowledgment our laboratory partners at the National Institute of Standards and Technology Rapid Drug Analysis and Research program (NIST RaDAR). We would also like to acknowledge our community partners at Bienestar Human Services, Community Health Project LA, The Sidewalk Project, and Homeless Outreach Program’s Integrated Care System (HOPICS). We would also like to extend our gratitude to the anonymous participants of Drug Checking Los Angeles who made this work possible through their donation of samples for confirmatory testing.

